# Xylazine Awareness, Desire, Use and Exposure: Preliminary Findings from the Rhode Island CUTS Drug Checking Cohort

**DOI:** 10.1101/2024.02.29.24303571

**Authors:** Ju Nyeong Park, Rachel Serafinski, Merci Ujeneza, Michelle McKenzie, Jessica Tardif, Alex J. Krotulski, Adina Badea, Elyse R. Grossman, Traci C. Green

## Abstract

**Background:** Xylazine, an α2 adrenergic receptor agonist, is a veterinary sedative that causes severe health complications yet interventions to detect, prevent and treat human exposure remain underdeveloped. Community-based drug checking services (DCS) involve the consensual collection and testing of small amounts of drugs to increase community awareness and reduce drug-related harms. This study characterized xylazine awareness, desire, use and exposure among people who use drugs (PWUD) in Rhode Island, USA.

**Methods:** We linked and analyzed DCS and survey data from an ongoing cohort of PWUD. Between February and August 2023, 125 PWUD were recruited and enrolled from harm reduction organizations and surveyed about xylazine awareness and use behaviors. Using point-of-care Fourier Transform infrared spectroscopy (FTIR-S), at least one drug sample was tested from each participant and confirmed offsite at a laboratory. Results were conveyed in real-time, along with harm reduction education, referrals to resources and care.

**Results:** Virtually all participants (99%) wanted to avoid xylazine exposure. Half (51%) knew what xylazine was, and a quarter (26%) suspected previous exposure. Xylazine exposure was primarily surmised through sedating (45%) and ulcerative (26%) effects. Only 9% of participants submitted a sample that they perceived to contain xylazine. Xylazine was detected in 14% of samples using FTIR-S and in 21% of samples using a dual laboratory approach of gas chromatography mass spectrometry (GC-MS) and liquid chromatography quadrupole-time-of-flight mass spectrometry (LC-QTOF-MS). Participants thought that these xylazine-positive samples were fentanyl (77%), heroin (14%), or Percocet® (9%).

**Conclusion:** Implementing point-of-care DCS at harm reduction organizations could be useful in rapidly increasing xylazine awareness and engaging at-risk individuals in prevention, harm reduction, treatment, and rapid care for xylazine-related wounds.

## Introduction

The opioid epidemic costs the US economy over one trillion dollars annually.^1^ Opioids, including fentanyl, heroin, and counterfeit opioid pills, account for two-thirds of overdose deaths.^2^ Xylazine—a veterinary sedative and an α2 adrenergic receptor agonist—has emerged as a health threat among people who use drugs (PWUD).^3,4,5^ Xylazine causes severe clinical effects including central nervous system depression and necrotizing skin/soft tissue infections.^6,7^ Xylazine-associated overdoses may require more extensive medical care than other overdoses.^8^ Additionally, xylazine-induced lesions occur both at injection and peripheral sites, and appear even if the drug is smoked or snorted. These wounds can be resistant to healing and lead to amputations.^9^ Between 2018-2021, xylazine increased by 1,238% among overdoses.^3^ Though data are limited, case reports and data are emerging on xylazine-associated morbidity and mortality.^4-7,10^

Federal support for fentanyl test strip (FTS) programs burgeoned from 2021 when federal agencies authorized the use of funding to distribute FTS. In Rhode Island (RI) alone, over 50,000 FTS were distributed in 2023. However, the implementation of comprehensive drug checking services (DCS) lags that of FTS programs. DCS involve the consensual collection and testing of small amounts of drugs including xylazine.^11,12,13^ Additionally, DCS can promote harm reduction behaviors and linkages to care among PWUD and help public drug surveillance efforts.^7,11,12,14,15,16,17,18^ There are more than 16 DCS in North America that have tested 49,786 samples.^12^ However, unlike fentanyl, the study of xylazine detection, prevention, and treatment is relatively new. Accordingly, we sought to understand xylazine awareness, use, and exposure among a preliminary cohort of PWUD from RI.

## Methods

### i. Setting

The Community Use and Testing Study (CUTS) is an 18-month prospective cohort study of 600 RI participants that combines point-of-care DCS with biannual surveys. Enrollment of the first 125 participants occurred through community outreach and word-of-mouth between February and August 2023; recruitment locations included harm reduction organizations, housing services, and public spaces where overdoses occur. Eligible participants underwent informed consent. The interviewer-administered baseline survey (Qualtrics, Provo UT) took 45-60 minutes. Participants were compensated $40. The study was approved by the Lifespan Institutional Review Board and developed in consultation with the COBRE on Opioid and Overdose Community Advisory Board.^19^

### ii. Participants

Eligible participants were ≥18 years; spoke and understood English; used an illicit drug in the past 30 days; RI residents; and provided ≥1 sample for testing. Eligible samples included drug packaging containing remnant solid drug (i.e., baggie, wax fold); or a once-used cooker, cotton, or straw, and excluded storage containers, syringes, pipes, and used crack stems due to potential signal interference (e.g., contamination from reuse).

### iii. Survey measures

The survey contained previously-developed measures^20,21,22^ and included: (1) socio-demographics (e.g., age, sex, gender identity, race/ethnicity, primary language, education, employment, housing); (2) medical co-morbidities; (3) overdose; (4) drug treatment; (5) FTS use; (6) drug use and social network characteristics; and (7) xylazine awareness, concerns, and perceived exposure, among other measures.

### iv. Drug checking

Remnant drug samples were collected from participants throughout the study and tested by trained staff in community spaces using Fourier Transform infrared spectroscopy (FTIR-S) and FTS, then sent for laboratory-based confirmatory testing. At the time of enrollment, FTS were available through local service organizations but xylazine test strips/kits were not being distributed in RI.

Our DCS protocols were based on two North American DCS.^23,24^ The entire process took 15-20 minutes. First, samples were scraped onto a sterilized scanning plate and scanned via the FTIR-S. Staff cleaned the FTIR-S between each tested sample using isopropyl alcohol. Next, they transferred the sample into a disposable 1 oz cup, and diluted as described below for testing with FTS. The remaining sample in its original packaging was secured in a mylar envelope and transferred to a laboratory. Any untested samples and packaging were discarded using a drug-neutralizing disposal bag.

#### a. Immunoassay-based fentanyl test strips (FTS)

Staff placed dissolved each sample with 5 mL of sterile water. For samples suspected to be methamphetamine (crystal or powder) or Adderall, the solution was further diluted to 30 mL of water due to concerns of false positives, and testing was repeated. The Rapid Response FTS (BTNX, Pickering, Ontario) was placed into the solution for ten seconds and read after 5 minutes.

#### b. Fourier-transform infrared spectroscopy (FTIR-S)

FTIR-S (Bruker Alpha Inc., Billerica, Massachusetts) was previously validated for use.^25,26^ FTIR-S can rapidly determine multiple active and inactive components and their relative proportion to one another. We adapted the testing protocol outlined in a program in British Columbia^24^ and the technician accessed trainings and technical support from the MADDS team.^25^

Trained technicians examined generated spectra and compared data to known spectra in library databases (e.g., Bruker pharmaceutical libraries, the Science Working Group for the Analysis of Seized Drugs library, the British Columbia Centre on Substance Use library, and the TICTAC library). A spectrum for xylazine is contained in the latter three libraries.

Staff explained limitations of FTIR-S and FTS before conveying results to the participant. These limitations included: 1) drug checking does not provide a guarantee of safety; 2) drug checking does not provide evidence of purity or dose; 3) people respond differently to drugs and drug checking does not provide personalized information about how you or anyone else will respond; 4) the information you receive is not an endorsement of a drug or of how a drug is used and is provided for the purpose of reducing harm; and 5) the FTIR-S and FTS may occasionally miss fentanyl, fentanyl analogues, or other dangerous substances such as xylazine. Participants also received the limitations of the results and disclaimers in writing during the informed consent process.

Preliminary results included the chemical components detected, including active and inactive cuts. When these results were verbally communicated to participants at the time of testing, the team also provided harm reduction education and information on the services available, including referrals to local medical and harm reduction organizations (e.g., wound care guidance and kits), further information from regional DCS (e.g., StreetCheck bulletins), and reinforcement on the use of extant harm reduction tools (e.g., how to use FTS).

#### c. Laboratory testing

The first sample collected per participant was transported to the Center for Forensic Science Research and Education (CFSRE, Horsham, PA) for confirmatory testing. Testing was conducted using combined qualitative and quantitative methods (when mass was sufficient). The laboratory used an Agilent Technologies (Santa Clara, CA) gas chromatograph mass spectrometer (GC-MS) for qualitative and quantitative analysis, and a SCIEX (Framingham, MA) liquid chromatograph quadrupole time-of-flight mass spectrometer (LC-QTOF-MS) for qualitative analysis. Samples were aliquoted, weighed (quantitative only), and prepared by a basic liquid-liquid extraction for GC-MS analysis, and subsequent mobile phase dilution for LC-QTOF-MS analysis. Datafiles were acquired in a non-targeted fashion to detect the presence of all relevant components with processing against an extensive in-house library database containing more than 1,200 targets. Only results confirmable through verification concurrent with standard reference materials were reported.

Qualitative results were reported in parts (e.g., fentanyl 1p, xylazine 5p, 4-ANPP 0.1p), where the primary drug was set to 1p and all other components were determined based on peak area ratio to the primary drug. Quantitative results were reported in percent composition (e.g., fentanyl 10%, xylazine 50%. 4-ANPP 1%) compared to the total mass taken for analysis (e.g., fentanyl 10% = 0.3 mg fentanyl of 3 mg total weight). Quantitation was performed via an external calibration model with internal standard comparing instrument response of the samples to known responses generated by analysis of standard reference materials at increasing increments. Both assays were validated prior to use and quality controlled within batch.

#### d. Sample-based questions and communication of confirmatory results

StreetCheck is an open-source platform designed to standardize and support the expansion of DCS created by MADDS in collaboration with community partners.^27^ It is an efficient, secure, and flexible environment for collecting and managing DCS data. The platform allows for follow-up questions and relies upon anonymous numeric and QR sample codes for tracking sample entry, analysis, and reporting. StreetCheck also contains detailed and standardized pharmacological and medical information on detected chemicals that is relayed back to participants.

Staff entered the following data into StreetCheck: date of collection, a photo of the substance, suspected substance(s), whether it was consumed and, if so, any reactions. They also conversed with each participant to obtain valuable information about their experience with the drug. This information was crucial to provide context to the sample analysis and offered more personalized and in-depth messaging when communicating the results. The public-facing website (streetcheck.org) organized sample-level data by sample ID and locality, and provided aggregate trends. Each participant received a unique anonymous weblink for each sample. Participants could also request their results from staff during visits.

### v. Data Analysis

Analysis was accomplished by merging the baseline survey and DCS data using a common sample ID. Using Stata/MP Version 16 (StataCorp, TX), descriptive characteristics were calculated. Cohen’s Kappa coefficient was used to measure the pairwise concordance between various detection methods (perceived, FTIR, laboratory). The Kappa coefficient ranges from -1 to +1 and a score of 0.4-0.59 indicates weak agreement, 0.6-0.79 indicates moderate positive agreement, and >0.8 indicates strong agreement between the variables.^28^

## Results

Of the preliminary cohort (N=125), 55% were male, and median age was 40 years. The cohort was racially and ethnically diverse (**Table 1**). Most completed high school (64%). Only 22% had stable housing. Most reported using cocaine (92%), fentanyl (67%), heroin (66%), and/or methamphetamine (46%) in the past 6 months. Half (54%) had a history of overdose and survived a median of 4 overdoses. Most participants carried naloxone (81%); some reported current receipt of methadone (55%) or buprenorphine (28%) treatment.

**Table 1:**
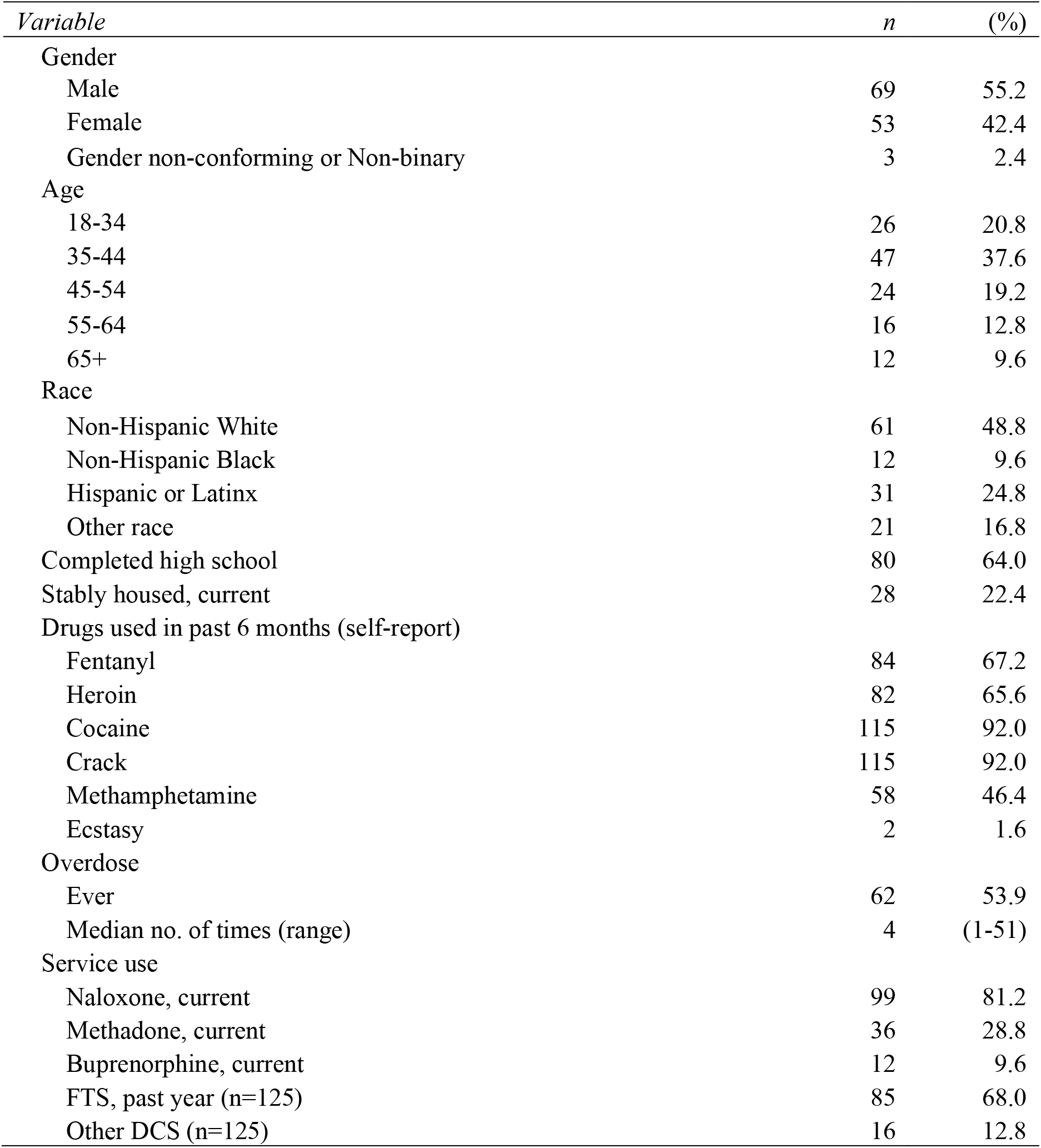
Baseline demographic, drug use and service use characteristics of the Community Use and Testing Study (CUTS) Cohort, Rhode Island (N=125)

A variety of samples were submitted for testing; most samples were perceived to contain crack cocaine (48%) or fentanyl (32%) with few submitting what they perceived to be xylazine (9%), heroin (7%) and methamphetamine (4%). Participants rarely submitted non-medical prescription opioids (e.g., Percocet) and powder cocaine (2%). Most (70%) had used the drug prior to submission.

Xylazine desire, knowledge, and experiences varied substantially (**Table 2**). Half (51%) knew what xylazine was and less than 1% wanted xylazine. A quarter (26%) suspected previous exposure. In contrast, 53% of the sample wanted fentanyl (data not shown). At baseline, xylazine exposure was primarily deduced from use experience (e.g., through its sedating (45%) and ulcerative (26%) effects) rather than known by individuals prior to its use (e.g., drug testing kits, DCS, urine testing, communications from their supplier).

**Table 2:**
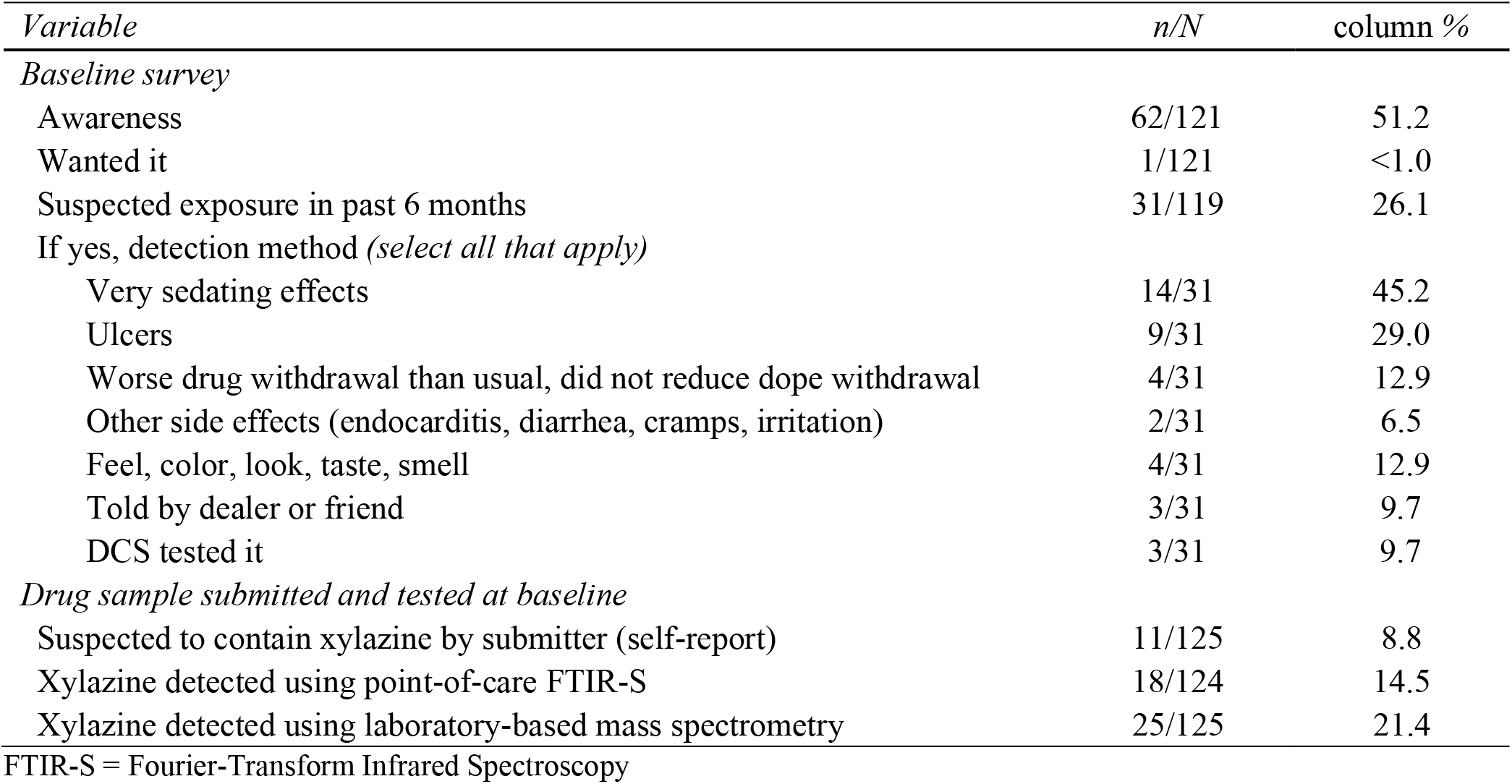
Xylazine desire, knowledge and experiences among the Community Use and Testing Study (CUTS) cohort, Rhode Island (N=125)

| *Variable* | *n/N* | *column %* |
| --- | --- | --- |
| *Baseline survey* |  |  |
| Awareness | 62/121 | 51.2 |
| Wanted it | 1/121 | <1.0 |
| Suspected exposure in past 6 months | 31/119 | 26.1 |
| If yes, detection method (*select all that apply*) |  |  |
| Very sedating effects | 14/31 | 45.2 |
| Ulcers | 9/31 | 29.0 |
| Worse drug withdrawal than usual, did not reduce dope withdrawal | 4/31 | 12.9 |
| Other side effects (endocarditis, diarrhea, cramps, irritation) | 2/31 | 6.5 |
| Feel, color, look, taste, smell | 4/31 | 12.9 |
| Told by dealer or friend | 3/31 | 9.7 |
| DCS tested it | 3/31 | 9.7 |
| *Drug sample submitted and tested at baseline* |  |  |
| Suspected to contain xylazine by submitter (self-report) | 11/125 | 8.8 |
| Xylazine detected using point-of-care FTIR-S | 18/124 | 14.5 |
| Xylazine detected using laboratory-based mass spectrometry | 25/125 | 21.4 |

FTIR-S = Fourier-Transform Infrared Spectroscopy

While only 9% of participants submitted a sample purportedly containing xylazine, it was detected in 14% of samples using FTIR-S and in 22% of samples using laboratory methods (**Table 2** and **Figure 1**). Eight samples were of sufficient size for xylazine quantification and contained 0.4%-11.8% xylazine (Table 3). Xylazine was detected among a range of other active substances, at both minor and trace levels. Xylazine was not detected in any samples without the presence of fentanyl. Xylazine-positive samples (n=23) were marketed to participants as fentanyl (78%), heroin (13%), or oxycodone.

**Table 3:** Unique samples submitted by Community Use and Testing Study (CUTS) participants that tested positive for xylazine in laboratory testing (N=25)

| Sample sold as... | Sample perceived to contain... | Psychoactive drugs detected using point-of-care FTIR | Psychoactive drugs detected using Laboratory-based mass spectrometry* | Percentage of Xylazine detected^ |
| --- | --- | --- | --- | --- |
| Fentanyl | Fentanyl, Methamphetamine | Fentanyl | Acetylfentanyl, Fentanyl, 4-ANPP, **Xylazine** | - |
| Heroin | Heroin, Fentanyl, **Xylazine** | Fentanyl, Caffeine, **Xylazine** | Fentanyl, Caffeine, 4-ANPP, Phenethyl-4-ANPP, **Xylazine** | - |
| M30 Pill (Percocet, Perc30) | Fentanyl | Sertraline | Sertraline, Fentanyl, **Xylazine** | 0.4% |
| M30 Pill (Percocet, Perc30) | Fentanyl | Sertraline | Sertraline, Fentanyl, **Xylazine** | - |
| Unknown | Fentanyl, **Xylazine** | Fentanyl, **Xylazine** | Acetylfentanyl, Fentanyl, 4-ANPP, **Xylazine**, Ethyl-4-ANPP, Phenethyl-4-ANPP | - |
| Fentanyl | Fentanyl | Fentanyl, **Xylazine** | **Xylazine**, Fentanyl, Phenethyl-4-ANPP, Acetylfentanyl, 4-ANPP, Ethyl-4-ANPP | 11.8% |
| Fentanyl | Fentanyl, **Xylazine** | Fentanyl, **Xylazine** | **Xylazine**, Fentanyl, Cocaine, Phenethyl-4-ANPP, 4-ANPP, Ethyl-4-ANPP | 4.3% |
| Fentanyl | Fentanyl, **Xylazine** | Fentanyl, **Xylazine** | Fentanyl, **Xylazine**, 4-ANPP, Phenethyl-4-ANPP | 5.6% |
| Fentanyl | Fentanyl, **Xylazine** | Fentanyl, **Xylazine** | Fentanyl, **Xylazine**, 4-ANPP, Phenethyl-4-ANPP | - |
| Fentanyl | Fentanyl | Fentanyl, **Xylazine** | **Xylazine**, Fentanyl, 4-ANPP | - |
| Fentanyl | Fentanyl, **Xylazine** | Fentanyl, **Xylazine** | **Xylazine**, Fentanyl, Cocaine, 4-ANPP, Phenethyl-4-ANPP | - |
| Fentanyl | Fentanyl | Fentanyl, **Xylazine** | **Xylazine**, Fentanyl, 4-ANPP | - |
| Fentanyl | Fentanyl, **Xylazine** | Fentanyl, **Xylazine** | **Xylazine**, Fentanyl, 4-ANPP | 6.8% |
| Fentanyl | Fentanyl | Fentanyl, **Xylazine** | **Xylazine**, Fentanyl, 4-ANPP, Phenethyl-4-ANPP | 6.5% |
| Unknown | Heroin | Fentanyl | Fentanyl, 4-ANPP, Acetylfentanyl, para-Fluorofentanyl, Heroin, Caffeine, Phenethyl-4-ANPP, Ethyl-4-ANPP, **Xylazine**, Lidocaine | - |
| Heroin | Heroin | Fentanyl, Lidocaine | Fentanyl, 4-ANPP, para-Fluorofentanyl, Lidocaine, Caffeine, **Xylazine** | - |
| Heroin | Heroin | Fentanyl, Lidocaine | Fentanyl, 4-ANPP, para-Fluorofentanyl, Caffeine, **Xylazine**, Lidocaine | - |
| Fentanyl | Fentanyl | Fentanyl, | Fentanyl, **Xylazine**, 4-ANPP, Caffeine | - |
| Fentanyl | Fentanyl, **Xylazine** | Fentanyl, **Xylazine** | Fentanyl, **Xylazine**, 4-ANPP, Cocaine, para-Fluorofentanyl, Acetylfentanyl, Phenethyl-4-ANPP, Caffeine, Ethyl-4-ANPP | 5.7% |
| Unknown | Fentanyl | Acetaminophen, Caffeine, Fentanyl, Lidocaine | Fentanyl, Acetylfentanyl, Caffeine, 4-ANPP, **Xylazine**, Acetaminophen, Cocaine, Phenethyl-4-ANPP, Ethyl-4-ANPP, Lidocaine | 3.8% |
| Fentanyl | Fentanyl | Fentanyl, Lactose | Fentanyl, **Xylazine**, 4-ANPP, Cocaine, Phenethyl-4-ANPP, Caffeine | - |
| Fentanyl | Fentanyl, **Xylazine** | Fentanyl, **Xylazine** | Fentanyl, **Xylazine**, 4-ANPP | - |
| Fentanyl | Fentanyl | Fentanyl | Fentanyl, 4-ANPP, Diphenhydramine, Cocaine, **Xylazine**, Phenethyl-4-ANPP | - |
| Fentanyl | Fentanyl | Fentanyl, **Xylazine**, Caffeine | Fentanyl, 4-ANPP, para-Fluorofentanyl, Cocaine, **Xylazine**, Caffeine, Phenethyl-4-ANPP, para-Fluoro-Phenethyl-4-ANPP, Diphenhydramine, Ethyl-4-ANPP | - |
| Fentanyl | Fentanyl | Fentanyl | Fentanyl, 4-ANPP, Cocaine Diphenhydramine, Phenethyl-4-ANPP, **Xylazine** | - |
| Fentanyl | Fentanyl | Fentanyl | Fentanyl, **Xylazine**, 4-ANPP, Phenethyl-4-ANPP | - |

\*laboratory results are reported in decreasing detection levels of active drug components.
^ If missing, mass was insufficient for quantification or quantification was unavailable at the time of sample processing

**Figure 1:**
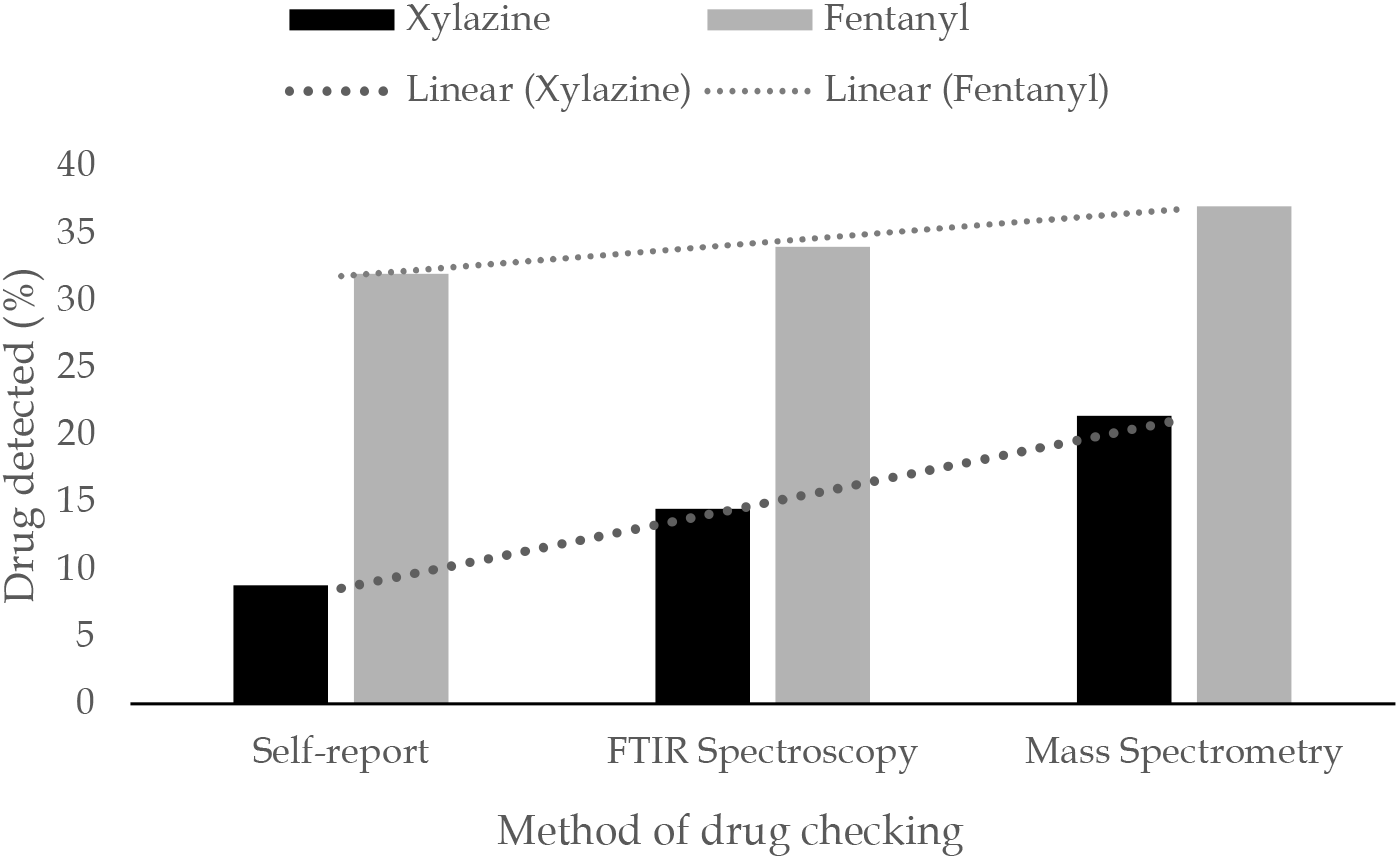
Xylazine and fentanyl detection using multiple methods among the first 125 Community Use and Testing Study (CUTS) cohort drug samples, Rhode Island. Note: The slope of the linear trend visually represents the differences between self-reported and laboratory-confirmed xylazine detection (black line) and fentanyl detection (grey line). Statistical agreement between self-reported and actual xylazine detection was weak (kappa<0.6). In contrast, the agreement observed between self-reported and actual fentanyl detection was strong (kappa >0.8).

Fentanyl detection was more consistent across the three methods (**Figure 1**). Whereas 32% perceived that their sample contained fentanyl, 34% and 37% of samples tested positive for fentanyl using FTIR-S and laboratory testing, respectively. Measure of concordance between perceived (i.e., self-reported) and laboratory-confirmed xylazine using Kappa statistic was 0.42 (Z=5.14; *p* < 0.001). For fentanyl, it rose to 0.85 (Z=9.24, *p* < 0.001). The concordance between perceived and FTIR-detected xylazine was 0.57 (Z=6.64, *p* < 0.001) and for fentanyl it rose to 0.78 (Z=8.71, *p* < 0.001); the concordance between FTIR-S and laboratory-confirmed xylazine was 0.74 (Z=8.18, *p* < 0.001) and for fentanyl it rose to 0.96 (Z=10.38, *p* < 0.001). However, in comparing the full range of other active drugs detected by the FTIR-S and laboratory to self-reported perceptions of the sample’s contents (**Table 3**), there was substantial variability and clear gaps in knowledge.

## Discussion

Although the earliest records of DCS in the US date back to the 1960s,^29^ point-of-care DCS are yet to be scaled up or rigorously examined in the US context. Our findings extend previous literature that has detected xylazine in drug supplies and documented its clinical effects.^5-10^ A DCS in Philadelphia recently detected xylazine in >90% of fentanyl/heroin samples with concentrations ranging from 5-70%.^30^ Xylazine awareness was moderate in our baseline cohort, a vulnerable population comprised of mostly unstably housed and polysubstance-using RI residents at risk of overdose. We found that in the absence of DCS, PWUD in RI relied on subjective health effects to decipher xylazine exposure as related to experiences of sedation and ulcerative wound appearance. Notably, the statistical agreement between what was perceived by PWUD and detected through laboratory testing was weak (kappa<0.6). In contrast, the agreement observed between perceived and actual fentanyl exposure was strong (kappa >0.8).

This highlights the potential for point-of-care DCS to fill knowledge gaps when newer drugs enter the illicit market by directly affirming and expanding the public’s awareness of local drug supplies. Point-of-care DCS provides a rich opportunity for learning from PWUD about their experiences with the drug sample, communicating results and providing access to harm reduction supplies.

In the absence of a regulatory framework for DCS, validation studies for xylazine tests will need to be conducted for rapid point-of-care testing tools. DCS models that include laboratory-based confirmation testing are advantageous over single-drug rapid tests as the former is more comprehensive and can be rapidly expanded to include novel psychoactive drugs as the drug supply evolves.

### Limitations

To our knowledge, this is one of few prospective studies in the US that have integrated DCS into research. However, we caution that the experiences in RI may not be generalizable outside of the state. The clinical and public health significance of trace amounts of xylazine detected in submitted samples remains unknown and will require further evaluation. The percentages of xylazine detected in this study cannot be interpreted as population-level prevalence estimates as non-random sampling was used to recruit participants and collect samples.

### Conclusions

In this DCS study, we detected substantial knowledge gaps regarding the composition of the local drug supply. Implementing DCS at harm reduction organizations could rapidly increase community awareness of xylazine and other contaminants, and engage PWUD into prevention and other critical supports such as wound care. Substantial federal and state investments for DCS implementation and research, as well as policy supports (e.g., drug paraphernalia law reform) will be required to scale up DCS across the country.^31,32^

## Data Availability

All data produced in the present work are contained in the manuscript

